# Reinforcement Learning for Chronic Care Pathway Optimization: A Unified Framework across Three Clinical Goal Types

**DOI:** 10.64898/2026.07.03.26357209

**Authors:** Ruilei Wang, Huifen Chen, Yifan Wu, Zhaoping Li, Ruiming Shen, Fengyao He, Shuai Zhao, Nansheng Zheng

## Abstract

Chronic care requires sequential treatment under competing biomarker, safety, and cost constraints, yet clinical goal structures differ across diseases. This version asks whether one physiology-informed reinforcement-learning paradigm can be written once and then instantiated for three goal structures— treat-to-target, multi-marker cruise, and bounded cycle completion—without a disease-specific policy architecture. Gout, chronic kidney disease, and PCOS-mediated fertility treatment are the three instances. Training is behaviour cloning followed by proximal policy optimization on 500 simulated trajectories per disease. Evaluation uses paired seeds (N = 50 primary; N = 500 bootstrap 95% confidence intervals), multi-seed offsets 42–51, ablation, and out-of-distribution profiles.

PCOS (cycle completion): PPO 72.0% against the doctor baseline 54.0% at N = 50; at N = 500, PPO 69.8% [65.6, 73.8] against doctor 52.8% [48.8, 57.2]. Gout (treat-to-target): PPO 88.0% against 90.0% (McNemar p = 1.0). CKD (cruise): doctor 32.0%, BC/PPO 38.0%.

This v2 withdraws three bibliographic coordinates that appeared in v1 and that do not match any recoverable source. The withdrawn comparison is replaced by a NOR-Gout calibration at weeks 4, 8 and 12 that belongs to the recalibrated kernel of the parallel journal manuscript. Shared BC→PPO training is instantiated across the three goal types without cross-disease weight sharing. Prospective validation remains necessary.

**Highlights:**

- Three goal types are hosted as different success tests, not as a claimed clinical taxonomy.
- PCOS cycle completion +18 percentage points against the doctor baseline at N = 50.
- Gout remains non-inferior to the guideline-mimicking doctor in the v1 evaluation table.
- NOR-Gout weeks 4/8/12 belong to the recalibrated kernel, not Section 3.
- Behaviour-cloning warm-start remains necessary under the gout kernel used here.

## 1. Introduction

Sequential treatment in chronic disease requires balancing biomarker targets, adverse events, and cost under safety constraints. Reinforcement learning offers a principled approach to policy search over multi-step treatment pathways, yet healthcare applications demand physiology-aware simulators, reproducible baselines, and transparent external-validity assessment.

Most clinical reinforcement-learning studies optimize a single disease endpoint without asking how evaluation metrics, reward design, and success criteria should change when the underlying clinical goal shifts from a level that must be reached and held, to a corridor that must be kept, to a cycle that must be completed. The grouping used here follows that time structure of the success test. It is not offered as a clinically recognised taxonomy. Gout urate-lowering therapy is the treat-to-target instance (ACR/EULAR). CKD management is the cruise instance (KDIGO). PCOS ovulation induction is the cycle-completion instance. Whether one training paradigm generalizes across all three remains the question of this preprint.

Electronic-health-record reinforcement learning faces confounding by indication, and online exploration in living patients raises ethical barriers. Physiology-grounded simulators provide a controlled comparison under known dynamics. They do not replace later patient-level evaluation.

## 2. Methods

For each disease: (1) generate N = 500 guideline-mimicking doctor trajectories; (2) train behaviour cloning for 100 epochs with class-balanced cross-entropy; (3) fine-tune proximal policy optimization for 3000 episodes initialized from the clone with GAE (λ = 0.95, γ = 0.99). Minimal DQN and offline CQL baselines share the evaluation harness.

Primary endpoint is disease-specific success: gout, consecutive in-health steps per ACR treat-to-target; CKD, a stable cruise window per KDIGO multi-target maintenance; fertility, ovulation/health per PCOS ovulation-induction guidance. Secondary endpoints are cumulative reward, treatment cost, steps, and side-effect scores. Multi-seed evaluation uses offsets 42–51 (N = 50 each). Large-sample analysis uses N = 500 with percentile bootstrap (2000 resamples). Pairwise tests are McNemar (binary success) and Wilcoxon signed-rank (continuous), with Benjamini–Hochberg FDR across 27 cross-disease comparisons (α = 0.05).

### 2.1 Literature calibration (v2)

v1 compared gout febuxostat-only trajectories to a 2017 citation that cannot be recovered as stated. That comparison, and the associated correlation and error figures, are withdrawn.

The external calibration that replaces the withdrawn comparison is against NOR-Gout (Uhlig, Karoliussen, Sexton, et al., 2021) at weeks 4, 8 and 12, and belongs to the recalibrated kernel reported in the parallel journal manuscript; it is not the kernel behind the v1 evaluation table of Section 3. Published week-4/8/12 means are 413, 371 and 341 µmol/L, with attainment 21.3%, 48.7% and 69.3%. Locked simulation on the same weeks is 419.69, 373.96 and 341.89 µmol/L, with attainment 16.8%, 42.4% and 63.2%. The allopurinol kernel is Wright, Duffull, Merriman, Dalbeth, Barclay and Stamp, *British Journal of Clinical Pharmacology* 81:277–289 (2016). Out-of-distribution profiles (eGFR 15–25, ALT > 120, sUA > 800 µmol/L, and acute-on-chronic flare combinations; n = 20 repeats each) are retained from v1.

The table below records calibration readings of the recalibrated kernel.

**Table.** NOR-Gout weeks 4, 8 and 12 versus the locked simulation (v2 replacement of the withdrawn external-anchor paragraph).

| Week | NOR-Gout mean ($\mu\text{mol/L}$) | NOR-Gout attainment | Locked simulation mean | Locked simulation attainment |
| --- | --- | --- | --- | --- |
| 4 | 413 | 21.3% | 419.69 | 16.8% |
| 8 | 371 | 48.7% | 373.96 | 42.4% |
| 12 | 341 | 69.3% | 341.89 | 63.2% |

## 3. Results

PCOS remains the primary efficacy signal of this preprint: PPO 72.0% against doctor 54.0% at N = 50; at N = 500, PPO 69.8% [65.6, 73.8] against doctor 52.8% [48.8, 57.2]. Gout: PPO non-inferior, 88.0% against 90.0% (McNemar p = 1.0). CKD: doctor 32.0%, BC/PPO 38.0%. Offline CQL 92.0% on gout trajectories. Removing behaviour-cloning warm-start collapsed gout PPO attainment from 88.0% to 0.0% in the archived ablation. The gout readings are the v1 evaluation table, produced by the earlier kernel, retained unchanged in this revision.

Large-sample point estimates (N = 500) remain: PCOS doctor 52.8% [48.8, 57.2], PPO 69.8% [65.6, 73.8]; gout doctor 82.6% [79.0, 85.8], BC 82.6% [79.0, 85.8], PPO 81.2% [77.6, 84.4]; CKD doctor 28.8% [24.8, 32.8], PPO 35.4% [31.4, 39.6]. Those gout figures are likewise the v1 evaluation table, produced by the earlier kernel, retained unchanged in this revision.

### 3.1 Ablation and literature calibration

Removing BC warm-start collapsed gout PPO attainment from 88.0% to 0.0%, confirming that initialization is essential under the kernel used here. Crystal-pool ablation did not reduce attainment at N = 50. Literature calibration for the recalibrated kernel is the NOR-Gout table in Section 2.1; the Section 3 readings are the v1 evaluation table, and the two kernels are distinct. The withdrawn v1 correlation and error figures are not restated.

### 3.2 Preliminary expert vignette

Three senior clinicians (one rheumatologist, one nephrologist, one reproductive endocrinologist) blindly rated 20 synthetic cases on a 1–5 Likert scale. Median acceptability for the PPO policy was comparable to the guideline doctor baseline (PCOS 4.3 vs 3.9, p = 0.08; gout 4.1 vs 4.2, p = 0.75). The panel is a pilot (the pre-registered protocol specifies 20 vignettes per disease).

## 4. Discussion

The three instances show different pictures. PCOS supports exploring bounded cycles. Gout cautions against reading a learned policy as a single verdict against a mature guideline: PPO matches the doctor on attainment while using a different cost and step profile. CKD cruise attainment is low across strategies. All results are in silico. Absence of electronic-medical-record validation is the primary limit on external validity.

## Limitations

Simulated doctor baselines approximate but do not replicate real clinician heterogeneity. The NOR-Gout table in Section 2.1 belongs to the recalibrated kernel of the parallel journal manuscript; residual trajectory error remains, and the module horizon is not a statement about the cohort’s 12-month follow-up. Expert vignette validation used a small blinded panel (N = 3 specialists, 20 pilot cases) and synthetic trajectories only. Out-of-distribution tests and CQL offline baselines cannot substitute for prospective cohort validation.

v1 reported three bibliographic coordinates that do not match any recoverable source. They are withdrawn in this version and are not used in the parallel journal manuscript.

## 5. Conclusion

A treat-to-target instance, a cruise instance and a cycle-completion instance can share one training stack without shared policy weights. PCOS remains the primary efficacy signal of this preprint. Gout remains non-inferior to the guideline-mimicking doctor in the v1 evaluation table. CKD remains a cruise-control stress test. Prospective electronic-medical-record validation remains necessary before deployment claims.

## Data Availability

All synthetic patient data generated for this study are produced by the open-sourced patient generator (src/clinical/patientgenerator.py) with fixed random seeds (global seed=42, patient generator seed=422024). The complete data generation parameters, drug-effect matrices, and natural drift parameters are specified in config/gout.py and environment/reward.py (release v1.1). Pretrained model weights (BC and PPO) are included in the repository. Experimental outputs are archived as compressed archives, parquet/jsonl, and related files in the data/output directory. No real patient data were used in this study.

https://gitee.com/ruileiwang/NaviTreat

